# Patients’ experience to inform decision-making and clinical follow-up: The example of total hip arthroplasty

**DOI:** 10.1101/2023.08.10.23293821

**Authors:** Anne Lübbeke, Stéphane Cullati, Christophe Baréa, Sophie Cole, Gianluca Fabiano, Alan Silman, Nils Gutacker, Thomas Agoritsas, Didier Hannouche, Rafael Pinedo-Villanueva

## Abstract

**Background:** The aim of this project was to enable patients and clinicians to benefit from previous patients’ experiences with total hip arthroplasty (THA) by seeking out patients’ views on what is important for them, leveraging registry data, and providing outcome information that is perceived as relevant, understandable, adapted to a specific patient’s profile, and readily available.

**Methods:** We created the information tool “Patients like me” in four steps. (1) The knowledge basis was the systematically collected detailed exposure and outcome information from the Geneva Arthroplasty Registry established 1996. (2) From the registry we randomly selected 275 patients about to undergo or having already undergone THA and asked them via interviews and a survey which benefits and harms associated with the operation and daily life with the prosthesis they perceived as most important. (3) The identified relevant data (39 predictor candidates, 15 outcomes) were evaluated using Conditional Inference Trees analysis to construct a classification algorithm for each of the 15 outcomes at three different time points/periods. Internal validity of the results was tested using bootstrapping. (4) The tool was designed by and pre-tested with patients over several iterations.

**Results:** Data from 6836 primary elective THAs operated between 1996 and 2019 were included. The trajectories for the 15 outcomes from the domains pain relief, activity improvement, complication (infection, dislocation, peri-prosthetic fracture) and what to expect in the future (revision surgery, need for contralateral hip replacement) over up to 20 years after surgery were presented for all patients and for specific patient profiles. The tool was adapted to various purposes including individual use, group sessions, patient-clinician interaction and surgeon information to complement the preoperative planning. The pre-test patients’ feedback to the tool was unanimously positive. They considered it interesting, clear, complete, and complementary to other information received.

**Conclusion:** The tool basead on a survey of patients’ perceived concerns and interests and the corresponding long-term data from a large institutional registry makes past patients’ experience accessible, understandable, and visible for today’s patients and their clinicians. It is a comprehensive illustration of trajectories of relevant outcomes from previous “Patients like me”. This principle and methodology can be applied in other medical fields.

## Background

A major implication of replacing a joint by an implant – in this case a total hip arthroplasty (THA) – is that the person will be living with the implant for the rest of his/her life. Patients are keen to be informed about the benefits and harms of the intervention and how these evolve over the long-term. Patients about to undergo THA would benefit from the experiences of people similar to them, who have already had the surgery and lived with the prosthesis. However, the systematic long-term documentation of outcomes remains the exception rather than the rule in clinical practice, thus limiting the structured knowledge patients and clinicians can gain from previous experience. Even when it is available, the knowledge gained is rarely shared with patients but stays with the clinician or is only disseminated through scientific publications. Patients’ information needs are broad, seeking information from the clinician, but also from interpersonal sources, especially people like themselves (e.g. family members, friends) and/or through information material (leaflets, internet, television, social media, print media) [1]. The latter is in general broad and not specific to the patient’s individual circumstances.

Uncertainty regarding the experience of disease, treatment, prognosis, and specific risks related to one’s general health status are associated with stress and high emotional pressure [2]. Knowing what to expect before surgery and be offered shared decision-making positively influences patients’ outcomes and satisfaction with care after arthroplasty [3,4]. This requires clinicians to individualize their explanations to each patient, or at least to groups of patients that share similar attributes. This is typically done with the help of prognostic tools such as clinical prediction scores, risk stratification tools, or risk calculators derived from analyses of large datasets (administrative data, electronic health records, registries or cohorts). The aim and ambition of these prognostic tools is to provide individualised predictions, on one or more key outcomes, typically short-term, to inform patients about the likely benefits and harms from surgery [5,6]. Prediction models have become popular in recent years to inform clinical practice [7]. However, these models have often failed to predict the outcomes of orthopaedic surgery [8,9], sometimes due to few numbers of events or missing data on outcomes or predictors in the available databases. In general, the clinical impact of prediction models remains unproven [10,11].

A second type of tools is focused on summarising the evidence. These are often disseminated in scientific publications, such as systematic reviews or clinical practice guidelines (e.g. clinical practice guidelines [12]), but can also come in the form of patient decision aids, either printed or through online platforms or applications (e.g. Magic App, RECOVER-E) [13,14,15]. Decision aids are designed to support shared decision making either by preparing patients to interact with clinicians, or to enhance the conversation during the clinical encounter [16]. Decision aids aim to present the evidence on benefits and harms, their uncertainties, as well as practical issues, in a format that is understandable and intuitive to patients [17,18,19].

None of the current tools for prediction or decision-making cover relevant benefits and harms over the short-, mid- and long-term after THA, after matching patients’ profiles, and report on their needs, interests, and concerns. The aim of this project was to produce an information tool to enable patients and clinicians to benefit more directly from previous patients’ experiences with THA. This was done by seeking out patients’ views on what is important for them, leveraging corresponding registry data and producing outcome information perceived as relevant, understandable, adapted to a specific patient’s profile, and readily available.

## Methods

### Registry data

Essential condition for the project was the existence of a dataset with the relevant predictor and outcome information over the long-term. The different steps undertaken for the tool creation are summarized in Figure 1.

**Figure 1.**
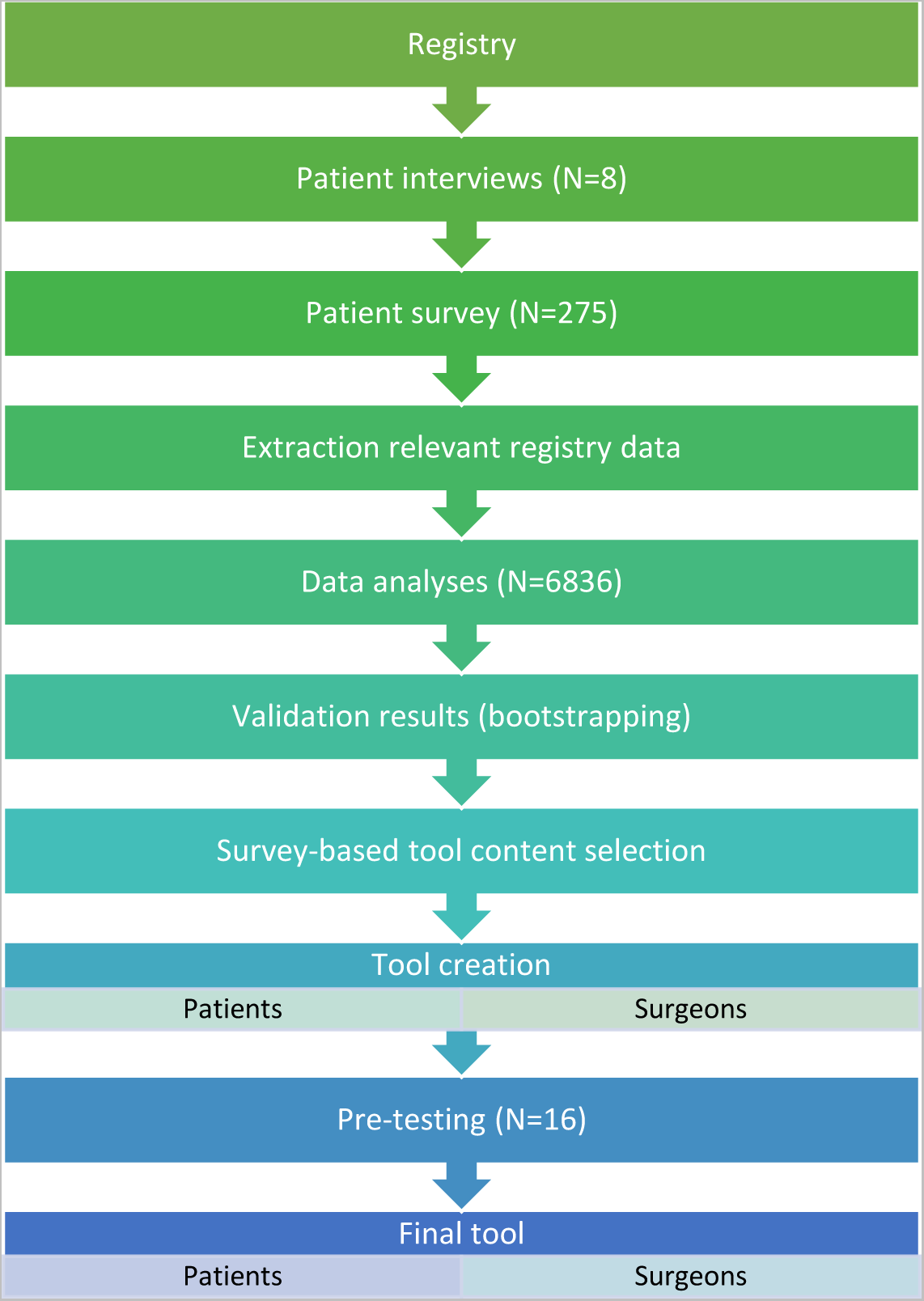
Building blocks of the project.

The Division of Orthopaedics at the Geneva University Hospitals established an institutional arthroplasty registry (GAR) in 1996. The institution is a large tertiary public hospital in a high-income country with universal health care coverage. The registry continuously and systematically collects detailed information about patients’ characteristics before surgery, about the surgery itself, and about the patients’ short-, mid-, and long-term experiences with their hip prosthesis. These data were the basis for the information tool. The registry has been described in detail elsewhere [20].

### Patient interviews and survey

To capture patients’ interests, needs, and concerns, we contacted 379 randomly selected patients from the registry either just about to undergo or having already undergone primary elective THA. Patients were invited to participate in a survey from 13 February to 17 September 2020. The questionnaire was specifically developed for this project and sent by mail to patients who were either just about to undergo surgery (n=95), or at 1 (n=94), 5 (n=95), or 10 years after surgery (n=95). We asked patients about their views on the benefits and harms associated with the operation and living their daily life with the prosthesis, and which they perceived as most important. The items of the questionnaire were chosen based on published literature, surgeons’ input (n=7), observations of patients attending a pre-operative course on THA, and one-to-one interviews with eight patients in December 2019 and January 2020. Ethics approval was obtained for use of the questionnaire within the registry and this project (N° CER PB_2017-00164).

### Registry data analysis

Outcomes of interest were selected based on survey participants’ responses and grouped into mutually exclusive categories. These were later mapped to specific questions included in GAR that could be used to measure outcomes in our patient population.

Patients who underwent a primary elective THA between March 1996 and December 2019 were included in the analysis. The end of follow-up was December 31, 2020. We excluded participants who had a large head (>28 mm), metal-on-metal bearing or a bilateral operation on the same day. Conditional Inference Tree (CIT) analysis [21] was used to construct classification algorithms based on pre-operative characteristics that identify clusters of patients with homogeneous outcomes. This analytical approach was used to differentiate between profiles of patients. Separate algorithms were developed for each relevant outcome and time point (i.e. at 1, 5, and 10 years after THA). Additionally, survival trees were generated using the classification and regression tree method to produce cluster-specific survival curves for outcomes reporting on clinical events that could happen at any time between registry follow-up points. CIT seeks to identify predictors that split the population into homogeneous subgroups (clusters) in terms of the variance in outcome. It does so by identifying variables that are of increasingly less importance to improving the classification until a point is reached when additional variables no longer have discriminatory power. The classification and regression tree method used to generate survival trees achieves the same objective but by identifying the pre-operative predictors that generate distinct survival curves.

A classification method was chosen over prediction models because the latter produce coefficients that are generally difficult for patients to understand, whereas classification is a natural concept that can be more easily discussed with patients in a clinical consultation. In addition, prediction models normally generate a probability that an individual will experience a given event (e.g. a fracture) or an expected result for a given outcome (e.g. mean WOMAC score), which even if the model is highly accurate means that, from the perspective of the patient, they will have to both understand the concept of probability and assess the number for the information to be meaningfully informative. Many patients will not meet those conditions. The classification approach, on the other hand, presents patients with a simpler view about how other people like them did (e.g. how many had fractures and how many didn’t, or the full distribution of their WOMAC scores) which is likely to add valuable information for patients.

Candidate predictors were selected from the GAR based on clinical input and evidence reported in published literature [22]. To mitigate the impact of missing data on results, imputation methods [23] were used to predict values for both outcomes and predictors.

To assess internal validity, 1000 bootstrap samples of equal size to the original sample were generated and the classification analysis undertaken again for each. As the classification method does not produce predicted values, performance of the models cannot be assessed by comparing predicted to observed. Instead, the bootstrap method allows to evaluate whether a different make-up of the sample would lead to different classification trees, which was informative for internal validity. Statistically significant predictors from the primary analysis were hence compared to the frequency of predictors identified as statistically significant in the 1000 bootstrapped CITs. The number of terminal nodes in each tree of the primary analysis was also compared to the average number of terminal nodes across the corresponding bootstrapped trees. This was done for each outcome and corresponding period of analysis.

### Tool creation and pre-testing

The tool “Patients like me” was designed by a graphic designer, along with tailored feedback from the team, around the topics most relevant to patients and based on the findings from the primary analysis. The pre-test of the information tool was conducted from January to March 2022 by the sociologist with patients participating either at the pre-operative education session or the post-operative follow-up consultations. Sixteen patients agreed to participate in the pre-testing (10 women, 6 men). They tested the information tool either online or on paper according to their personal preference. During the pre-test period, we modified and retested the tool twice.

## Results

### Survey results

Of the 379 posted questionnaires, 275 were returned complete (72.6%) and 37 were incomplete (9.8%). Among the patients who returned the survey complete, 54.2% were women. Participants’ mean age was 70 years (±11.3, range 36-95). Educational achievement of patients was mandatory school 32.0%, secondary level 31.2%, and tertiary level 36.8% (missing n=22). Among the patients who responded, 30.9% were soon to have their surgery (N=85), 21.1% were at 1 year postoperative (n=58), 29.1% at 5 years (n=80), and 18.9% at 10 years after surgery (n=52).

Benefits perceived as most important (≥80%) included: pain relief (92.1%), independence in walking and moving (90.3%), and return to daily activities at home (85.1%), (Table 1). Respondents were also asked to order the three most important benefits. The first most important benefit was pain relief (78%). For the second and third, no one single benefit was selected but a variety of them: returning to my leisure activities (31%), returning to my daily activities at home (17%), and independence in walking and moving (17%) for the second choice; and returning to my leisure activities (18%), sleep again (17%), and stopping or reducing pain medication (13%) for the third choice. When stratifying the responses by time to surgery results were similar compared to those at all time points combined.

**Table 1.** Importance of perceived benefits of hip replacement (n=275 patients’ responses)

|  | Important or very important | Mean* (SD) |
| --- | --- | --- |
| Pain relief | 92.1% | 4.6 (0.9) |
| Independence in walking and moving | 90.3% | 4.6 (0.8) |
| Return to my daily activities at home | 85.1% | 4.4 (1.1) |
| Resuming my social life (meeting friends, family) | 79.7% | 4.3 (1.2) |
| Return to my leisure activities (sport, travel, etc.) | 78.9% | 4.2 (1.3) |
| Stopping or reducing pain medication | 72.0% | 3.9 (1.4) |
| Emotional well-being | 71.1% | 4.0 (1.3) |
| Return to my professional activities (Not applicable=162 respondents) | 69.2% | 3.8 (1.5) |
| Sleep again | 66.7% | 3.9 (1.4) |

\*Answer categories ranged from 1 = not at all important to 5 = very important

The harms perceived as most important (≥80%) included: infection of my prosthesis (83%), persistent pain (81%), fracture of the bone surrounding my prosthesis (81%), loss of control over my health (80%), fracture of my prosthesis (80%), and dislocation of my prosthesis (80%) (Table 2). Patients were asked to order the three most important harms. No single one was selected for the first, second and third most important harms. However, a variety were selected: for the first most important, pain that spreads to other joints (42%) followed by fracture of the bone surrounding my prosthesis (12%); for the second most important, fracture of the bone surrounding my prosthesis (23%), inability to resume all my activities (13%), persistent pain (11%); and for the third most important, difference in length (13%), inability to resume all my activities (12%), fracture of the bone surrounding my prosthesis (11%). Of note, fracture of the bone surrounding my prosthesis was on top of the list across the three most important risks. When stratifying by time to surgery, all harms were mostly rated as important or very important by all groups of patients. Some results were similar compared to patients taken altogether. The risk of early change of my prosthesis due to wear or loosening was important across all patients’ groups and ranged between 81% (before surgery) and 69% (at 1 and 5 years).

**Table 2.**
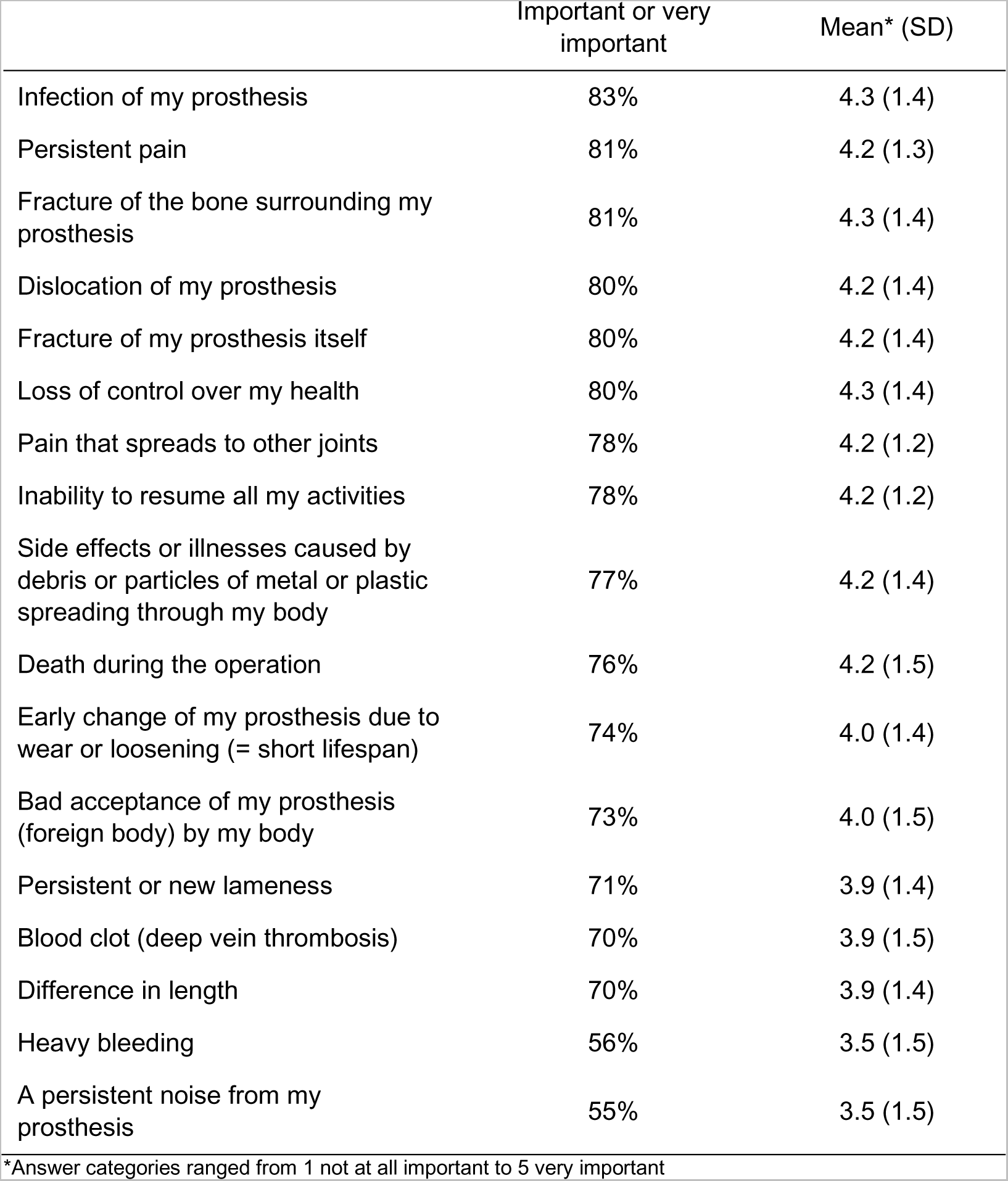
Importance of perceived harms of hip replacement (n=275 patients’ responses)

More generally, patients’ answers indicated pain relief, activity improvement, complications, and what to expect in the future as most important topics. Their views on what mattered to them remained consistent from before surgery to 10 years after surgery.

### Data analyses results

The most important benefits and harms of THA reported by patients were covered by one or more data items included in the GAR. The Western Ontario and McMaster Universities Arthritis Index (WOMAC) captures patients’ experience of pain and activity. The Short-Form 12 (SF-12), a patient-reported outcome measure assessing the impact of health on everyday life, includes questions about pain, independence, and interference. The UCLA Activity Scale (UCLA) assesses physical activity and includes a question about return to work and usual activities. The GAR also collect specific questions about pain medication as well as records of whether the patient experienced infection, fracture, dislocation, or a revision of their prosthesis. All benefits and harms were grouped into four outcome categories: pain, activity, complications, and expectations. Table 3 details the specific outcomes of interest as well as the corresponding questions/variable and the source used to measure them.

**Table 3.**
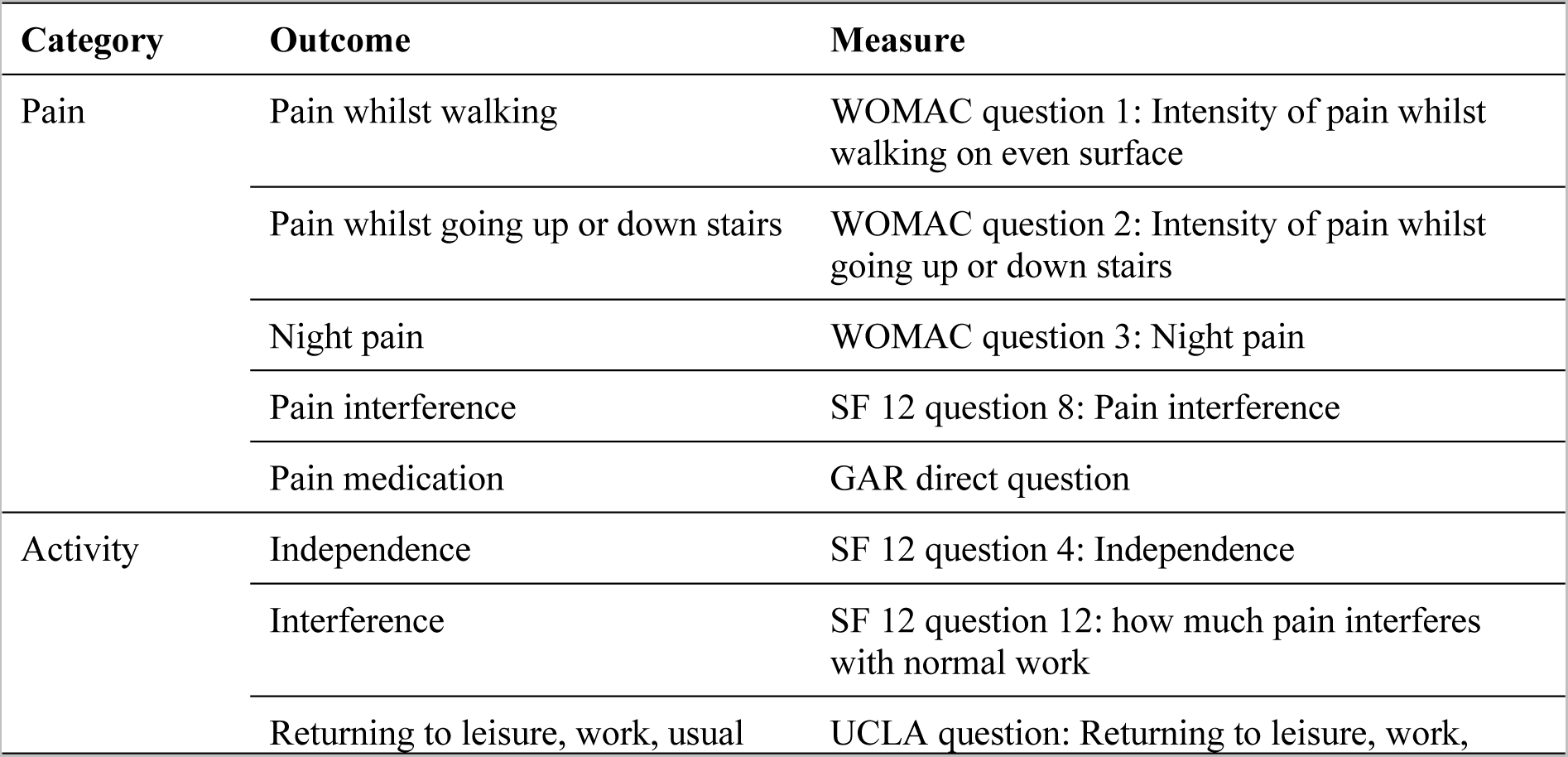
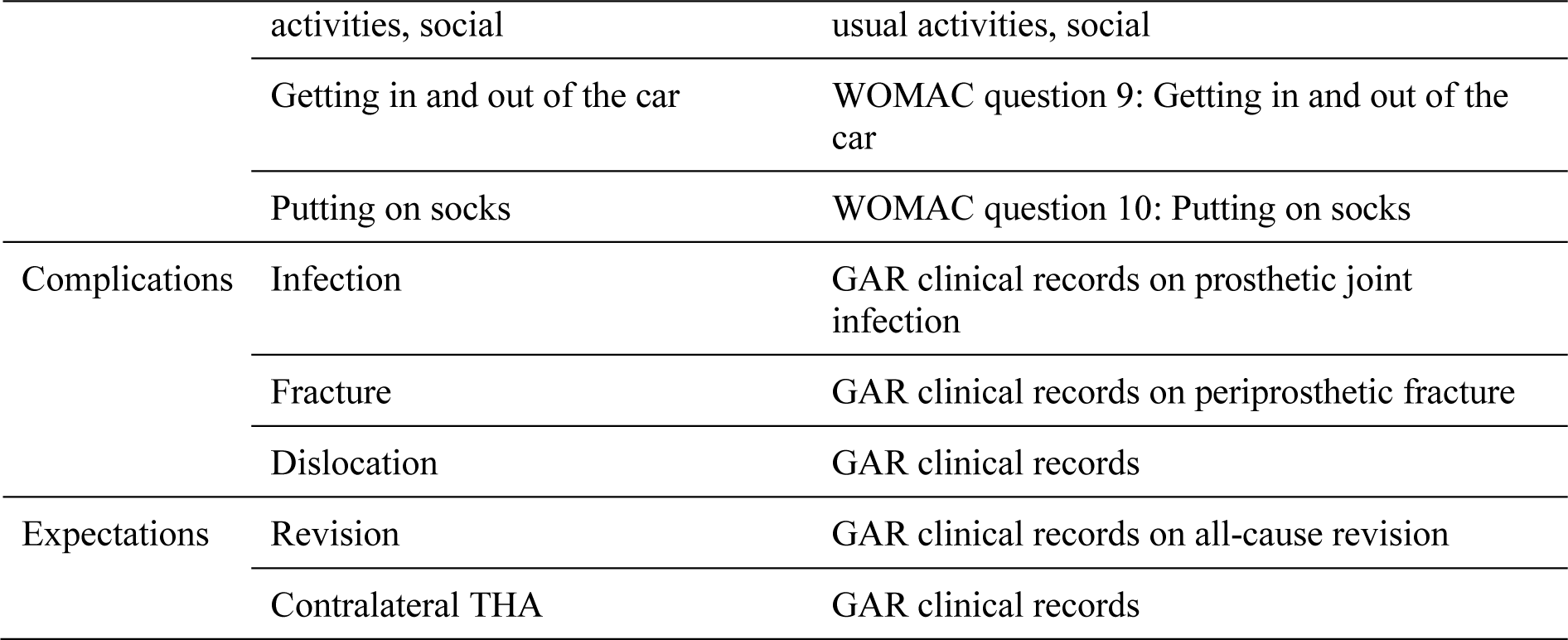
Outcomes of interest and corresponding measure/source.

Detailed results of the classification analysis for each outcome category are to be fully reported in upcoming manuscripts. A total of 6,836 operations were included in the CIT analysis. Characteristics of the patients are reported in Table 4. The final sample had more women (56.8%) than men and mean age was 68.9 (±12.2) years. Indication for surgery was mostly primary osteoarthritis (82%, n=5,610). Mean follow-up was 8.5 years (±5.7, range 0-24). Overall, 2,122 (31%) patients died between 1996 and 2020 and 347 (5.1%) were lost to follow-up.

**Table 4.**
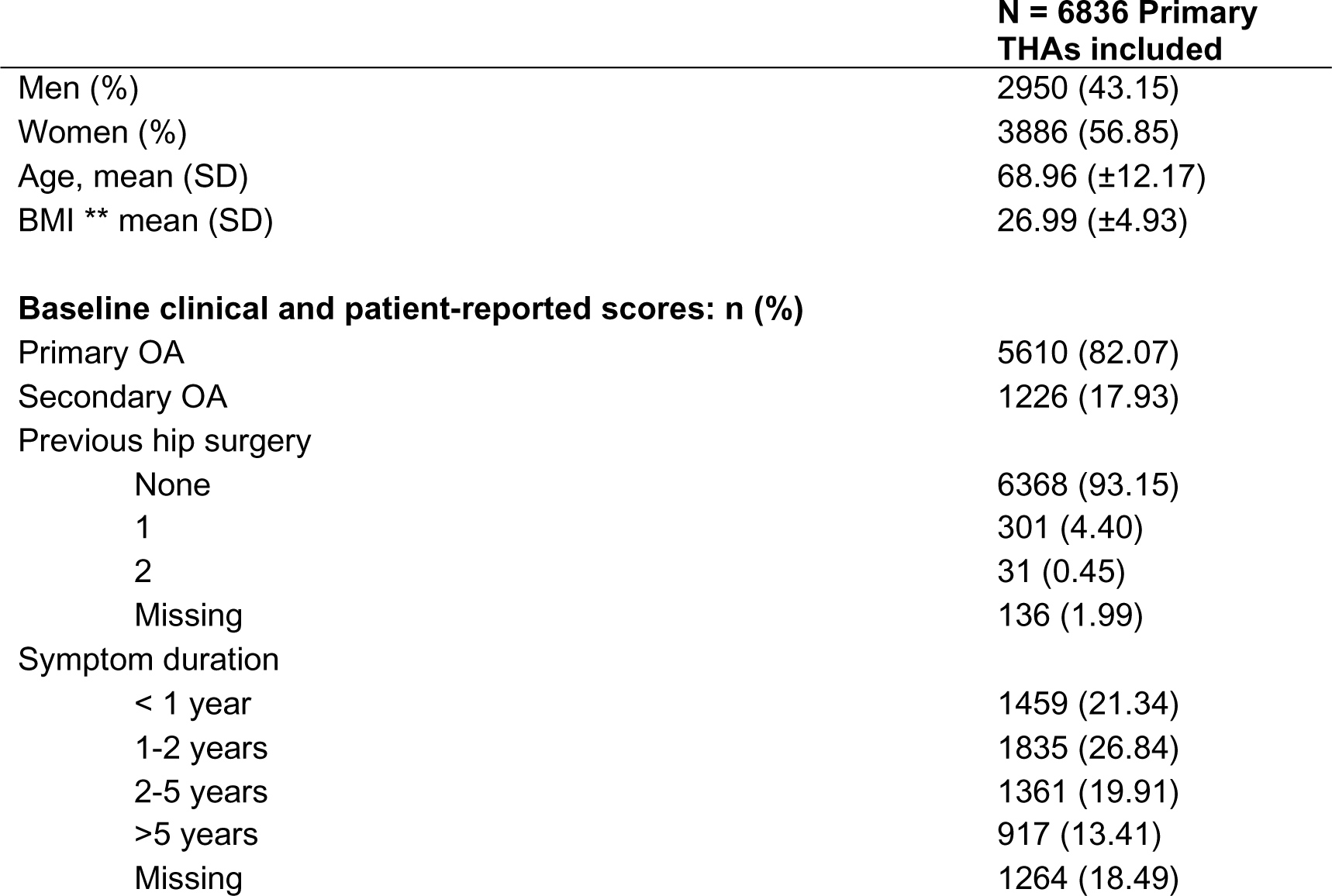
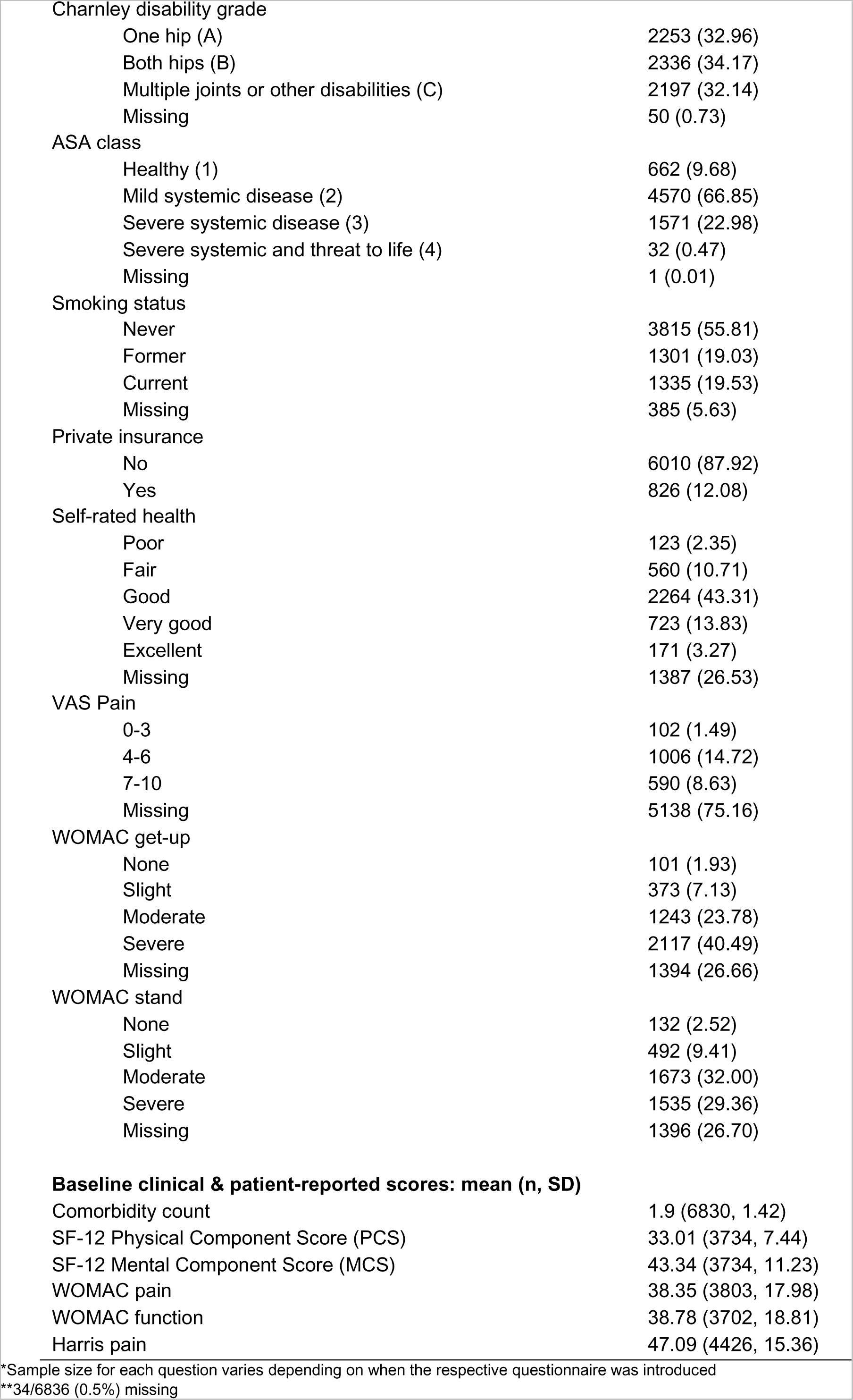
Baseline characteristics.

The CIT analysis was applied to all Pain and Activity outcomes at 1, 5, and 10 years after THA, as well as to all Complications and Expectations outcomes 1 year after surgery. Further, survival trees were generated for all Complications and Expectations outcomes up to 20 years after THA.

Candidate predictors were identified for each outcome; they included clinical and demographic variables such as age, sex, body mass index (BMI), comorbidity count, previous hip surgeries, underlying diagnosis, symptom duration, American Society of Anaesthesiologists (ASA) grade, Charnley disability grade, smoking status, and whether participants had public or private health insurance, for example. Other predictors included outcome variables measured at baseline such as specific WOMAC questions and overall scores, Harris pain and function scores, and specific SF-12 questions including self-rated health and the composite physical and mental component scores.

Missing data were more common with longer follow-up time. Reasons for not completing follow-up questionnaires included death, moving away from Switzerland, refusal to participate, and poor general health. Imputation methods were applied to predict values for missing data. There was no missing information on ASA grade, diagnosis, or insurance status.

Findings of the primary analysis are to be reported in figures of the resulting regression trees showing statistically significant predictors and threshold values, each generating a new branch into other predictors and thresholds, if relevant, until all cluster for the set outcome and time point were shown. Frequency distribution of the outcome variable will be included for each cluster.

### Outputs and testing

Three outputs were produced: (1) a 28-page information leaflet (web and print version) (Figures 2-5) for the patient intended for use during the patient-surgeon consultation at the time of the decision whether to operate or not, and prior to surgery as well as after surgery. The patient can read the leaflet on its own or discuss it with others. The tool can be used as support in a preoperative education session; (2) a digital visualisation for the surgeon (integrated in the registry) illustrating the patient’s profile and corresponding trajectories for all outcomes of past patients like her/him intended to complement the preoperative planning strategy; and (3) an 8-page infographic brochure summarizing the project’s approach, methods, results and applications intended to inform clinicians, researchers, and other health professionals from different specialties and institutions.

**Figure 2.**
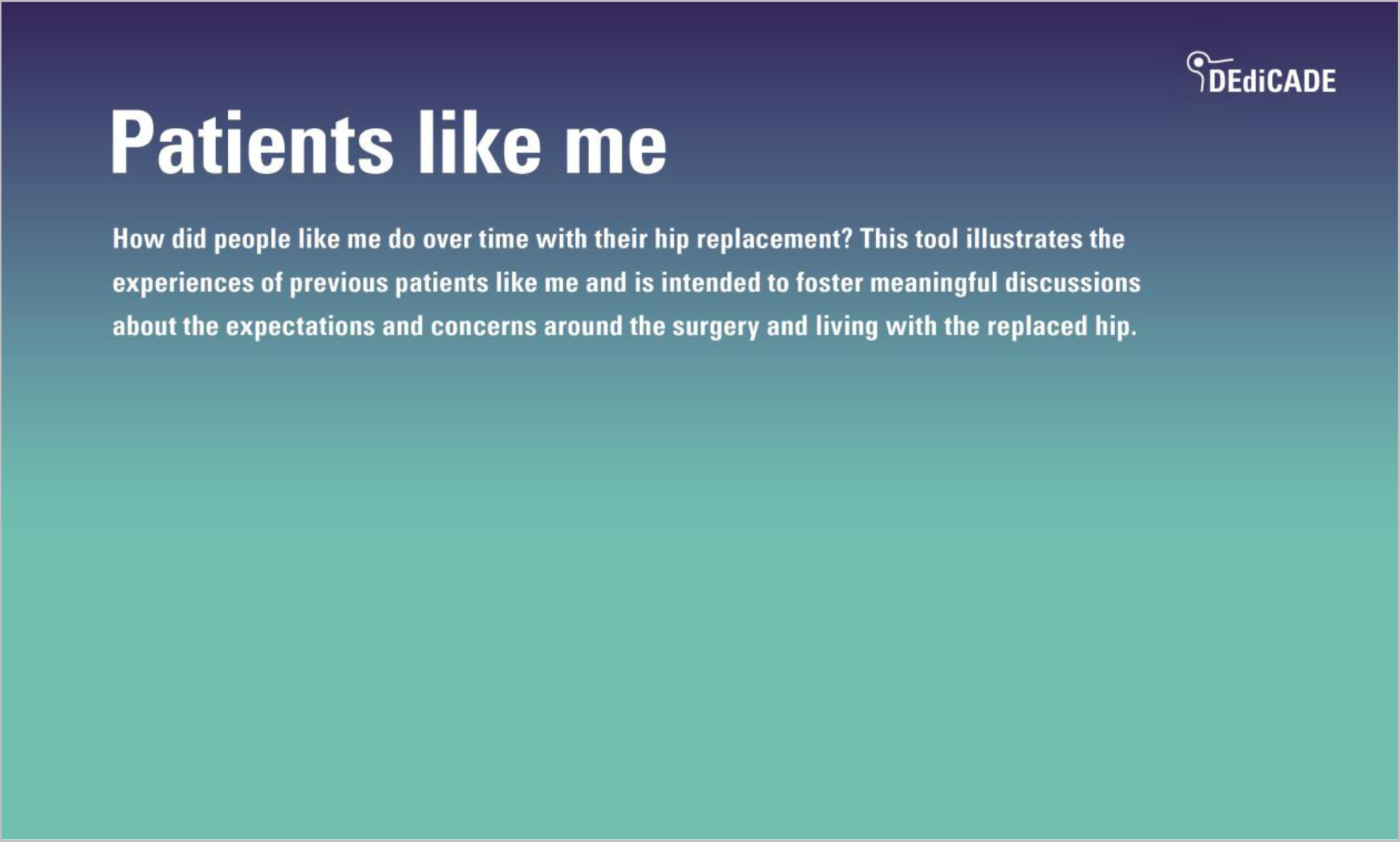
The patient information leaflet. (front page with image available upon request)

**Figure 3.**
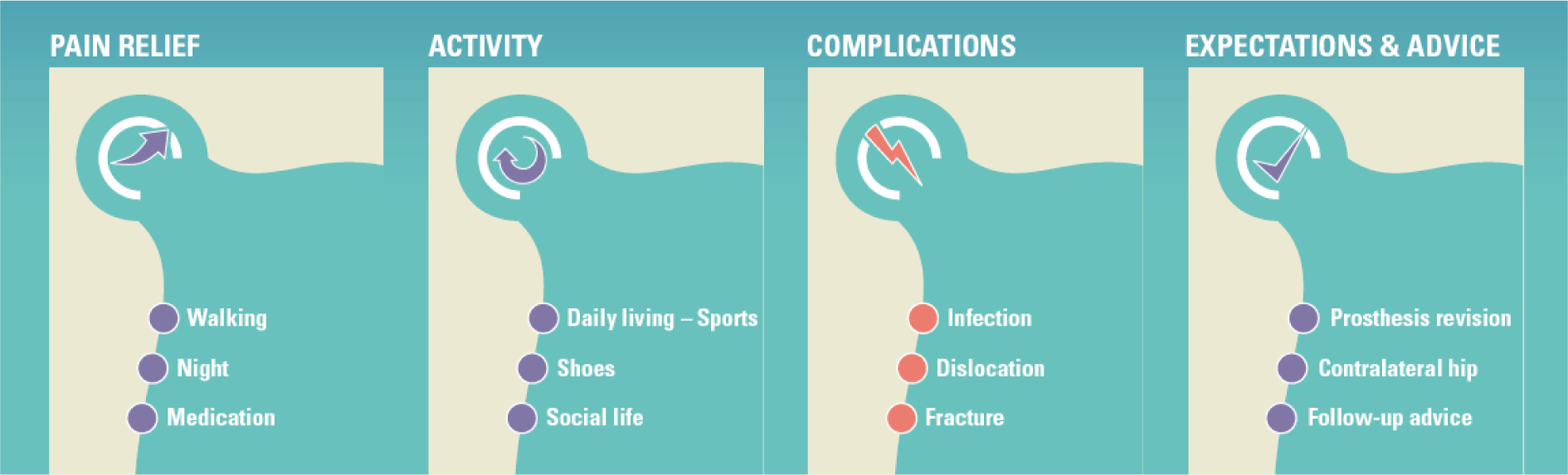
The four areas of interest to the patients.

**Figure 4.**
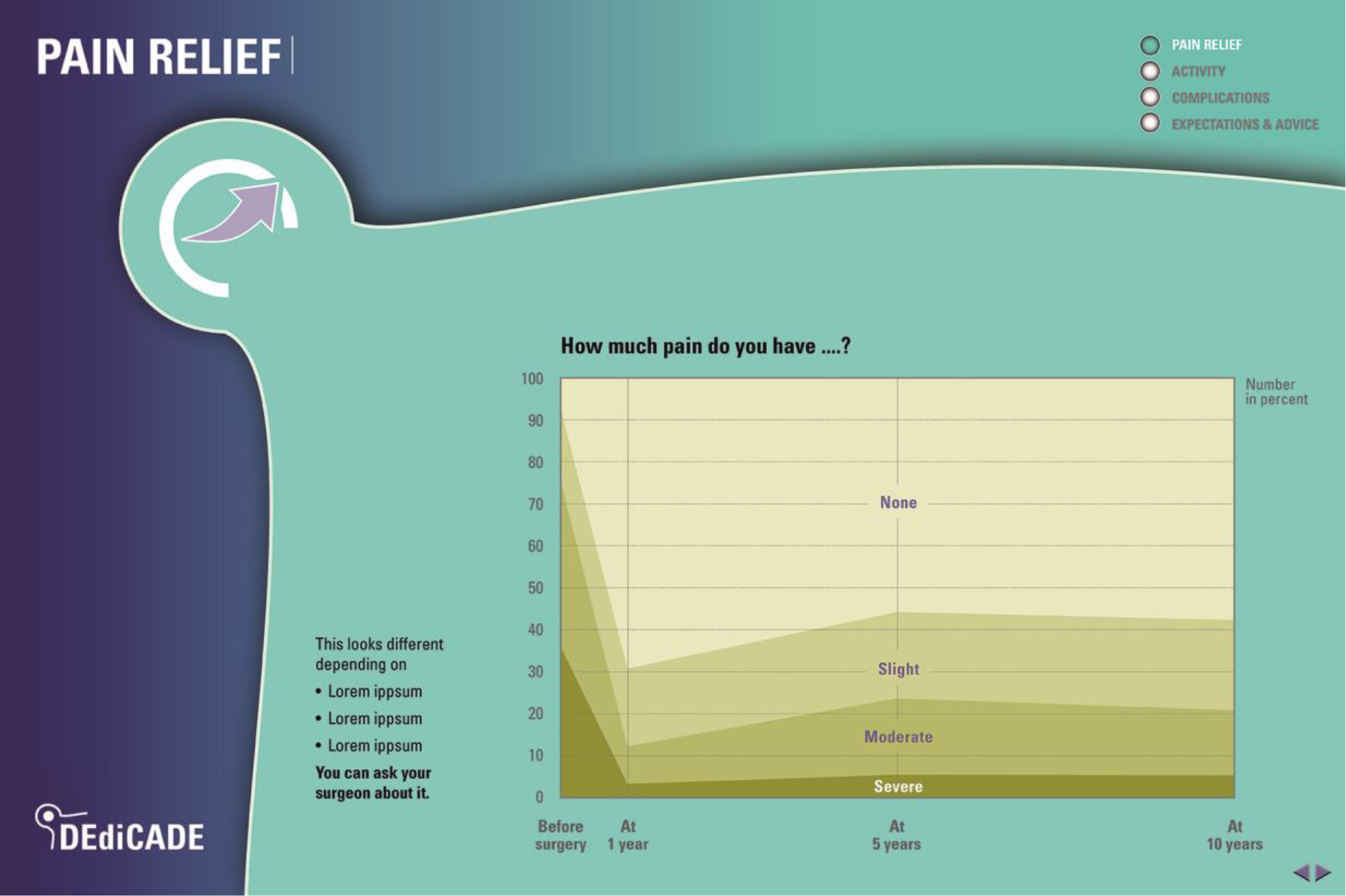
Pain during activity X before and after surgery as an example of symptom relief.

**Figure 5.**
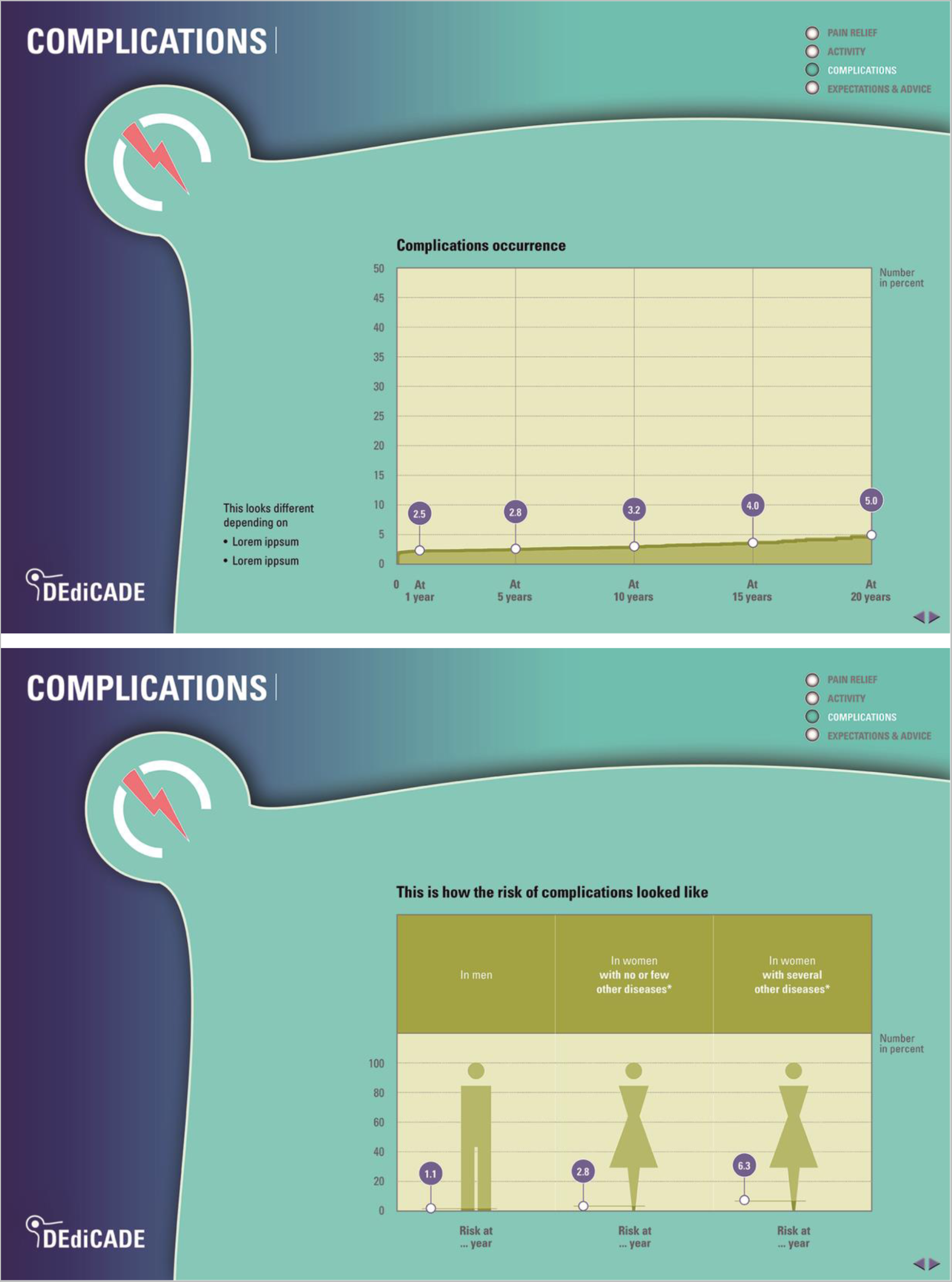
An example of a complication.

Pre-testing of the 28-page patient information leaflet showed that the feedback was positive across all patients. Regarding the content in the leaflet (information), patients appreciated the information, which was seen as interesting, clear and complete. The amount of information seemed a bit too much for few patients. Some appreciated that the information in the leaflet was complementary to that received from the surgeon during the patient-surgeon consultation, particularly on the issue of post-operative pain. First, the different feedbacks highlighted the need to clarify information on pain medication (i.e., that about 20% of patients report taking pain killers one year after surgery). A modified version containing more detail on pain medication was tested and there was no further criticism on this point. Regarding the form of the leaflet (colours, fonts, etc.), the overall impression was pleasant for all patients. The design and the colour were appreciated by most. A few patients suggested increasing the size of the smallest fonts, which was done. Pre-testing was undertaken either on a tablet or on the paper version if desired. Most patients were interested in taking home a paper version of the information tool.

## Discussion

In this project we have developed a comprehensive tool for patients and clinicians to discuss the entire care process of total hip replacement from prior to surgery to 20 years after surgery and to inform on multiple benefits and risks perceived as important by the patients. The information is tailored to groups (average outcomes) as well as to individual patients (outcomes provided by specific patient profiles). This project is an example of integration of registry-derived information into the clinical care process, and it highlights the importance/potential of clinical registries or databases in the learning health system to improve quality of care [24,25]. Past patients’ experiences were made accessible through the systematic documentation of the registry. The latter is an established, well-documented data collection infrastructure [26]. The pre-test patients’ feedback to the tool was unanimously positive. They considered it interesting, clear, complete, and complementary to other information received.

The material has been created for various circumstances and on different forms (web based, printed, integrated in the workflow via the registry). The implementation is immediate and inexpensive and has the potential to change the patient’s and clinician’s experience. We have shown here the case of THA, but using past patients’ experiences to inform clinical decision making and follow-up is obviously not limited to joint replacement.

Previous studies have used large datasets/registries to produce individualized predictions and inform patients e.g. about the likely quality of life benefit of surgery [27,28], or the risk of complications such as short-term revision and death [6], or both [29]. However, to our knowledge this is the first study that leveraged registry data to develop a comprehensive (meaning multiple harms and patient-reported benefits over the long-term) tool that allows to match a patient today to others (“Patients like me”) who had a THA in the past. It is also novel in that it uses clustering methods (via regression trees in our analysis) instead of clinical prediction or prognostic models to generate information about what surgery might bring about for patients and their clinicians. Prediction models can be and are indeed useful to guide clinical practice, but from the perspective of patients they are likely much less intuitively informative. Prediction models would result in an estimation of the likelihood that a specific patient experiences a particular outcome, commonly explained as a given number in 100 people having such fate [27,30], or more complicated yet as an odds ratio. These concepts are not straightforward or easy to understand for most. Although in our study we use regression models, we use them as a vehicle to identify variables to create clusters, which we can later match to the patient who is about to have surgery. By doing this, we can present prospective patients with information about 100 people like them, of whom not a predicted but a known number would have experienced the outcome of interest or those they want to avoid. We hypothesized that patients would understand it more easily, and their feedback suggests that they do. With this information easily accessible, patients can now be empowered to have more meaningful discussions with their clinicians, and ultimately improve the quality of care by improving the experience of shared decision-making and follow-up care.

Other initiatives have been launched recently with a similar aim of using routinely collected data from registries to inform personalised treatment. The DESTINY platform used by neurologists and psychiatrists in Germany helps to identify side effects or interactions of treatments, also collects patient data to build algorithms that can be used to predict treatment response [31,32]. Although predictions are not provided by patient clusters, the concept of using registry data to inform patient treatment follows a similar aim as our study.

The methods we employed were not without limitations. First, some variables reported high levels of missing data, which is a common problem of longitudinal datasets especially for patient-reported variables. To address this, we applied imputation methods, but these add some level of uncertainty to results. Also, when identifying clusters, the conditional tree approach we used finds thresholds in predictors using significance tests, which cannot be modified if greater levels of heterogeneity in the final clusters were preferable. This could be achieved using methods such as classification and regression tree (CART) analysis, however we opted for the conditional tree approach because it avoids the analyst having to decide on a number of parameters about the size and purity of nodes in the resulting tree, as well as having to manually prune it in the end, which can add significant bias.

The tool was derived from an institutional registry and the results may not be generalizable to other settings. However, baseline characteristics of the patients included in the registry are comparable to those of other national hip arthroplasty registries [26].

Although total hip replacement is already an established and highly successful intervention, there have been modifications in surgical practice - including implant selection and surgical techniques used – and minor changes in patient baseline status over the 20-year follow-up period. Consequently, the results from past patients presented in the information tool may not be exactly the same for today’s patients. To explore this, further analyses must be conducted with data from more recent years that will scarify length of follow-up but will allow examining the potential impacts of changes in practice.

## Conclusion

The information tool based on a survey of patients’ perceived concerns and interests and the corresponding long-term data from a large institutional registry makes past patients’ experience accessible, understandable, and visible for today’s patients and their clinicians. It provides information that is useful for them during the decision for surgery, and it offers profile-specific patient experience for meaningful discussions and expectation management perioperative and over the long-term. Finally, it provides tailored information complementing the surgeon’s preoperative planning strategy. Potentially complementing prediction models, the tool is a comprehensive illustration of trajectories of relevant outcomes up to 20 years after THA from previous “Patients like me”.

## Declarations

a. Competing interests: None to declare
b. Ethics approval: We confirm that all experiments were performed in accordance with relevant guidelines and regulations (such as the Declaration of Helsinki). Ethics approval for this project was obtained from the local ethics committee (Commission cantonale d’éthique de la recherche de Genève (CCER), n° PB_2017-00164). All necessary patient/participant consent has been obtained and the appropriate forms archived.
c. Funding: The DEdiCADE project was funded by the “Fondation privée des HUG”, www.fondationhug.org/en.
d. Availability of data and materials: The datasets generated and/or analysed during the current study are not publicly available due to local data protection rules, but de-identified data are available from the corresponding author on reasonable request and after permission from the local ethics committee.
e. Authors’ contributions: A.L., S.C., A.S., N.G., T.A., D.H. and R.P. designed the study. R.P., S.C., C.B., S.C., G.F. and N.G. performed or supervised the analyses. A.L., S.C., T.A. and R.P. interpreted the results. A.L. wrote the first draft of the manuscript. All authors contributed to the editing and revision of the manuscript.

## Data Availability

Availability of data and materials: The datasets generated and/or analysed during the current study are not publicly available due to local data protection rules, but de-identified data are available from the corresponding author on reasonable request and after permission from the local ethics committee.

## f. Acknowledgements

This project would not have been possible without the outstanding work of the graphic designer M. Uwe Otte. We would like to thank all the patients who have contributed information about their experiences before and after surgery through questionnaires and feedback at the clinical visits. We also thank all the orthopaedic surgeons and the registry staff who have contributed to the Geneva Arthroplasty Registry since 1996. Finally, we would like to thank Prof. Pierre Hoffmeyer and the « Fondation pour la recherche ostéo-articulaire » for the continuous support of the registry.

